# Bridging Genetics and Precision Medicine in Parkinson’s Disease through GP2

**DOI:** 10.64898/2026.03.26.26349418

**Authors:** Kajsa Atterling Brolin, Lara M. Lange, Emily Navarro-Jones, Simona Jasaityte, Yuan Ye Beh, Zih-Hua Fang, Hirotaka Iwaki, Lietsel Jones, Christine Klein, Teresa Kleinz, Hampton L. Leonard, Ignacio F. Mata, Alastair J Noyce, Njideka Okubadejo, Paula Saffie Awad, Laurel Screven, Ai Huey Tan, Marco Toffoli, Dan Vitale, Andrew Singleton, Cornelis Blauwendraat, Mike Nalls, Huw R Morris, the Global Parkinson’s Genetics Program (GP2)

## Abstract

In the Global Parkinson’s Genetics Program (GP2) we aim to advance precision medicine by integrating large-scale clinico-genetic data from diverse populations worldwide. We investigated potentially trial-eligible carriers of pathogenic and high-risk *GBA1* and *LRRK2* variants and conducted a global precision-medicine survey across GP2 sites. Among 65,509 individuals with Parkinson’s disease, we identified 9,019 (13.8%) potentially trial-eligible genetic variant carriers, including 6,789 *GBA1*, 2,084 *LRRK2*, and 146 dual *GBA1-LRRK2* carriers. Individuals were distributed across multiple global regions, many of which currently lack active gene-targeted trials, highlighting a global disparity between relevant variant carriers and the availability of disease modifying treatment trials. GP2’s unified framework supports equitable recruitment for gene-targeted therapeutic studies and helps address critical gaps in Parkinson’s disease genetics and future therapeutic development.

## INTRODUCTION

The increasing number of ongoing and planned clinical trials focusing on genetic forms of Parkinson’s disease (PD) reflects growing recognition of the critical role genetics plays in the pathogenesis of the disease. Genetic discoveries have been instrumental in guiding the development of targeted therapies for PD. For example, variants in *LRRK2*, the most common known cause of autosomal dominant PD, are believed to act through a gain-of-function mechanism, leading to increased kinase activity. Accordingly, several clinical trials are underway testing LRRK2 kinase inhibition and LRRK2 knock-down therapies. (*1, 2*) Similarly, *GBA1,* variability, which is linked to an increased risk for PD, encodes the lysosomal enzyme glucocerebrosidase, whose activity is typically reduced in variant carriers. This has prompted the development of trials aimed at enhancing glucocerebrosidase activity, reducing the accumulation of glucocerebrosidase substrates, or restoring lysosomal function. (*3, 4*) Previous studies, unrelated to PD, have shown that drugs supported by genetic evidence are 2.6 times more likely to pass a trial phase and have a higher success rate compared to those without (*5*), underscoring the importance of genetics. Despite these advances, the lack of disease-modifying treatments and the difficulty of recruiting ancestrally diverse, well-characterized participants for precision medicine initiatives remain challenges for many biotech and pharmaceutical companies.

The Global Parkinson’s Genetics Program (GP2; https://gp2.org) (*6*) is a global research initiative dedicated to addressing the evolving needs in PD genetics research, currently representing a consortium of 319 cohorts and clinical studies worldwide, including individuals with PD and Parkinsonism, individuals in prodromal PD stages, as well as healthy controls. By integrating clinico-genetic data from >250,000 participants of diverse ancestries worldwide, GP2 is positioned to accelerate precision medicine efforts in PD. As part of this initiative, we aim to leverage GP2’s extensive and in-depth genetic characterization of individuals with PD to inform recruitment strategies and to define a large, multi-ancestry, potentially trial-eligible cohort of gene variant carriers, thereby accelerating the translation of genetic discoveries into precision medicine. In this study, we aim to identify potentially trial-eligible *LRRK2* and *GBA1* variant carriers within GP2, summarize their clinical characteristics, and evaluate site-level readiness for genetically informed clinical trials.

## RESULTS

### Identification of trial-eligible gene variant carriers

Out of 65,509 unique individuals with PD, we identified a total of 9,019 (13.8%) potentially trial-eligible variant carriers, including 6,789 individuals carrying *GBA1* variants (6789/65509; 10.4%), 2,084 individuals with pathogenic or risk variants in *LRRK2* (2084/65509; 3.2%), and 146 dual *GBA1-LRRK2* carriers.

About 4.5% (303/6789) of *GBA1* variant carriers were known variant carriers, i.e., submitted as part of genetically enriched cohorts, while the remaining were newly identified *GBA1* variant carriers. The most frequent *GBA1* variants overall were p.E365K and rs3115534-G, followed by p.T408M and p.N409S. The p.N409S variant is known to cause Gaucher’s disease (GD) when in the biallelic state, while the other three variants have only been associated with an increased risk for PD. Other pathogenic variants linked to GD and PD risk were rarer; a detailed overview of all identified variants and the number of carriers is provided in Supplementary Table 1. Information on disease duration was available for 5471/6789 *GBA1* variant carriers (80.6%); of those, 2,034 (37.2%) had a duration of ≤3, 2,954 (54.0%) ≤5, and 3,628 (66.3%) ≤7 years. Extended clinical data were available for 2,062 carriers (30.4%). Regarding PD disease severity, the H&Y stage was available for 992/6789 (14.6%), including 606 (61.1%) with a H&Y stage ≤2 and 904 (91.1%) with a H&Y stage ≤3. Finally, data on cognitive status were available for 780/6789 (11.5%) participants; 458 (58.7%) of whom had normal cognition (defined as MoCA score ≥26/30 points), while 228 (29.2%) had mild cognitive impairment (defined as MoCA score ≥20 and <26 points).

About 22.3% (464/2084) of *LRRK2* variant carriers were known variant carriers from genetically enriched cohorts. The pathogenic p.G2019S variant was the most frequent *LRRK2* variant in the GP2 cohort overall, followed by the Asian risk variants p.R1628P and p.G2385R; a a detailed overview of all identified variants and the number of carriers is provided in Supplementary Table 1. Information on disease duration was available for 1,690/2084 (81.1%) *LRRK2* variant carriers; 471 (27.9%) had a disease duration of ≤3, 704 (41.7%) ≤5, and 926 (54.8%) ≤7 years. For 711 carriers (34.1%), extended clinical data were available. Data on disease severity were provided for 360 participants (17.3%); of those, 236 (65.6%) had a H&Y stage ≤2, and 323 (89.7%) a H&Y stage ≤3. 350 *LRRK2* carriers (16.8%) had information on cognitive testing; 198 (56.6%) of them had normal cognition, while 113 (32.3%) had mild cognitive impairment (MoCA score ≥20 and <26 points).

Finally, 146 individuals (0.2%) were dual carriers, harboring both a *GBA1* and a *LRRK2* variant. Detailed numbers on clinical data availability for single *GBA1* and *LRRK2* variant carriers as well as for dual carriers, including a breakdown by ancestry, are provided in **Table 1**.

**Table 1.** Overview of identified *GBA1* and *LRRK2* carriers per ancestry and corresponding clinical data.

| Ancestry | N carriers | N carriers with clinical data | H&Y (N) | H&Y≤2 (N) | H&Y≤3 (N) | MoCA (N) | MoCA ≥20 (N) | MoCA ≥26 (N) | Disease duration (N) | Disease duration ≤3 years (N) | Disease duration ≤5 years (N) | Disease duration ≤7 years (N) |
| --- | --- | --- | --- | --- | --- | --- | --- | --- | --- | --- | --- | --- |
| ***GBA1*** |  |  |  |  |  |  |  |  |  |  |  |  |
| **AAC** | 197 | 106 | 52 | 34 | 43 | 32 | 23 | 9 | 145 | 46 | 73 | 96 |
| **AFR** | 1485 | 156 | 58 | 30 | 51 | 24 | 19 | 8 | 1362 | 707 | 934 | 1058 |
| **AJ** | 469 | 284 | 108 | 64 | 93 | 95 | 83 | 61 | 429 | 133 | 218 | 267 |
| **AMR** | 165 | 39 | 21 | 10 | 19 | 22 | 17 | 12 | 84 | 17 | 28 | 40 |
| **CAH** | 146 | 64 | 48 | 37 | 47 | 35 | 28 | 12 | 116 | 32 | 55 | 73 |
| **CAS** | 76 | 7 | 2 | 2 | 2 | 1 | 1 | 1 | 30 | 6 | 9 | 21 |
| **EAS** | 192 | 85 | 28 | 17 | 25 | 31 | 26 | 13 | 127 | 44 | 63 | 82 |
| **EUR** | 3420 | 1233 | 659 | 403 | 608 | 472 | 427 | 307 | 2705 | 904 | 1334 | 1687 |
| **FIN** | 27 | 9 | 2 | 0 | 2 | 4 | 4 | 4 | 16 | 1 | 2 | 4 |
| **MDE** | 67 | 13 | 2 | 2 | 2 | 3 | 3 | 2 | 39 | 12 | 14 | 23 |
| **SAS** | 42 | 18 | 12 | 7 | 12 | 14 | 13 | 5 | 39 | 14 | 23 | 27 |
| **Other\*** | 503 | 48 | 0 | 0 | 0 | 47 | 42 | 24 | 379 | 118 | 192 | 250 |
| **TOTAL** | **6789** | **2062** | **992** | **606** | **904** | **780** | **686** | **458** | **5471** | **2034** | **2945** | **3628** |
| ***LRRK2*** |  |  |  |  |  |  |  |  |  |  |  |  |
| **AFR** | 3 | 0 | 0 | 0 | 0 | 0 | 0 | 0 | 3 | 1 | 2 | 2 |
| **AJ** | 465 | 228 | 121 | 89 | 110 | 115 | 108 | 75 | 379 | 111 | 157 | 211 |
| **AMR** | 60 | 16 | 13 | 5 | 10 | 10 | 9 | 5 | 26 | 8 | 10 | 15 |
| **CAH** | 32 | 9 | 5 | 3 | 5 | 0 | 0 | 0 | 24 | 9 | 14 | 14 |
| **CAS** | 24 | 3 | 1 | 1 | 1 | 2 | 2 | 1 | 11 | 4 | 5 | 8 |
| **EAS** | 598 | 142 | 97 | 58 | 81 | 26 | 25 | 15 | 482 | 152 | 223 | 290 |
| **EUR** | 633 | 280 | 117 | 74 | 110 | 173 | 147 | 84 | 547 | 127 | 214 | 273 |
| **MDE** | 79 | 17 | 6 | 6 | 6 | 8 | 4 | 3 | 72 | 13 | 21 | 31 |
| **SAS** | 1 | 0 | 0 | 0 | 0 | 0 | 0 | 0 | 1 | 1 | 1 | 1 |
| **Other\*** | 189 | 16 | 0 | 0 | 0 | 16 | 16 | 15 | 145 | 45 | 57 | 81 |
| **TOTAL** | **2084** | **711** | **360** | **236** | **323** | **350** | **311** | **198** | **1690** | **471** | **704** | **926** |
| **Dual carriers** |  |  |  |  |  |  |  |  |  |  |  |  |
| **AAC** | 1 | 0 | 0 | 0 | 0 | 0 | 0 | 0 | 1 | 0 | 0 | 1 |
| **AFR** | 2 | 1 | 0 | 0 | 0 | 0 | 0 | 0 | 1 | 1 | 1 | 1 |
| **AJ** | 54 | 25 | 7 | 4 | 7 | 9 | 9 | 8 | 40 | 7 | 13 | 16 |
| **AMR** | 2 | 1 | 1 | 0 | 1 | 1 | 1 | 0 | 2 | 1 | 1 | 2 |
| **CAH** | 3 | 1 | 1 | 1 | 1 | 1 | 1 | 0 | 2 | 0 | 0 | 0 |
| **CAS** | 1 | 0 | 0 | 0 | 0 | 0 | 0 | 0 | 1 | 0 | 1 | 1 |
| **EAS** | 20 | 8 | 3 | 2 | 3 | 0 | 0 | 0 | 15 | 5 | 6 | 8 |
| **EUR** | 43 | 19 | 8 | 6 | 6 | 5 | 4 | 3 | 36 | 8 | 14 | 17 |
| **MDE** | 8 | 1 | 1 | 0 | 1 | 1 | 1 | 1 | 8 | 0 | 2 | 3 |
| **Other\*** | 12 | 3 | 0 | 0 | 0 | 3 | 3 | 3 | 8 | 4 | 6 | 7 |
| **TOTAL** | **146** | **59** | **21** | **13** | **19** | **20** | **19** | **15** | **114** | **26** | **44** | **56** |

This table summarizes ancestry-stratified counts of individuals carrying *GBA1* variants, *LRRK2* variants, or both a *GBA1* and *LRRK2* variant (dual carriers). Columns indicate the number of variant carriers with available data for each clinical value and those meeting predefined thresholds for Hoehn and Yahr (H&Y) stage, Montreal Cognitive Assessment (MoCA), and disease duration.
AAC = African Admixed, AFR = African, AJ = Ashkenazi Jewish, AMR = Latinos and Indigenous People of the Americas, CAH = Complex Admixture, CAS = Central Asian, EAS = East Asian, EUR = European, FIN = Finnish, MDE = Middle Eastern, SAS = South Asian
\*Other reflects individuals for whom genetic ancestry could not be determined.

### Precision medicine survey

The results of our survey are summarized in **Fig. 1 and 2**. Among 153 GP2 sites contacted, 54 (35.3%) sites from 26 different countries across six continents responded (**Fig. 1**). Over half of them (53.7%) reported current or prior participation in movement disorder-focused clinical trials, while 27.8% had not yet participated but possessed the required infrastructure and interest, and 11.1% expressed interest in future participation pending appropriate infrastructure development (**Fig. 2A**). Most sites (94.4%) reported active participant recruitment for PD-related studies, including both interventional and observational studies (**Fig. 2B**). Nearly all responding centers indicated interest in being connected with academic or industry partners to facilitate recruitment of trial-eligible participants for future treatment trials.

**Figure 1.**
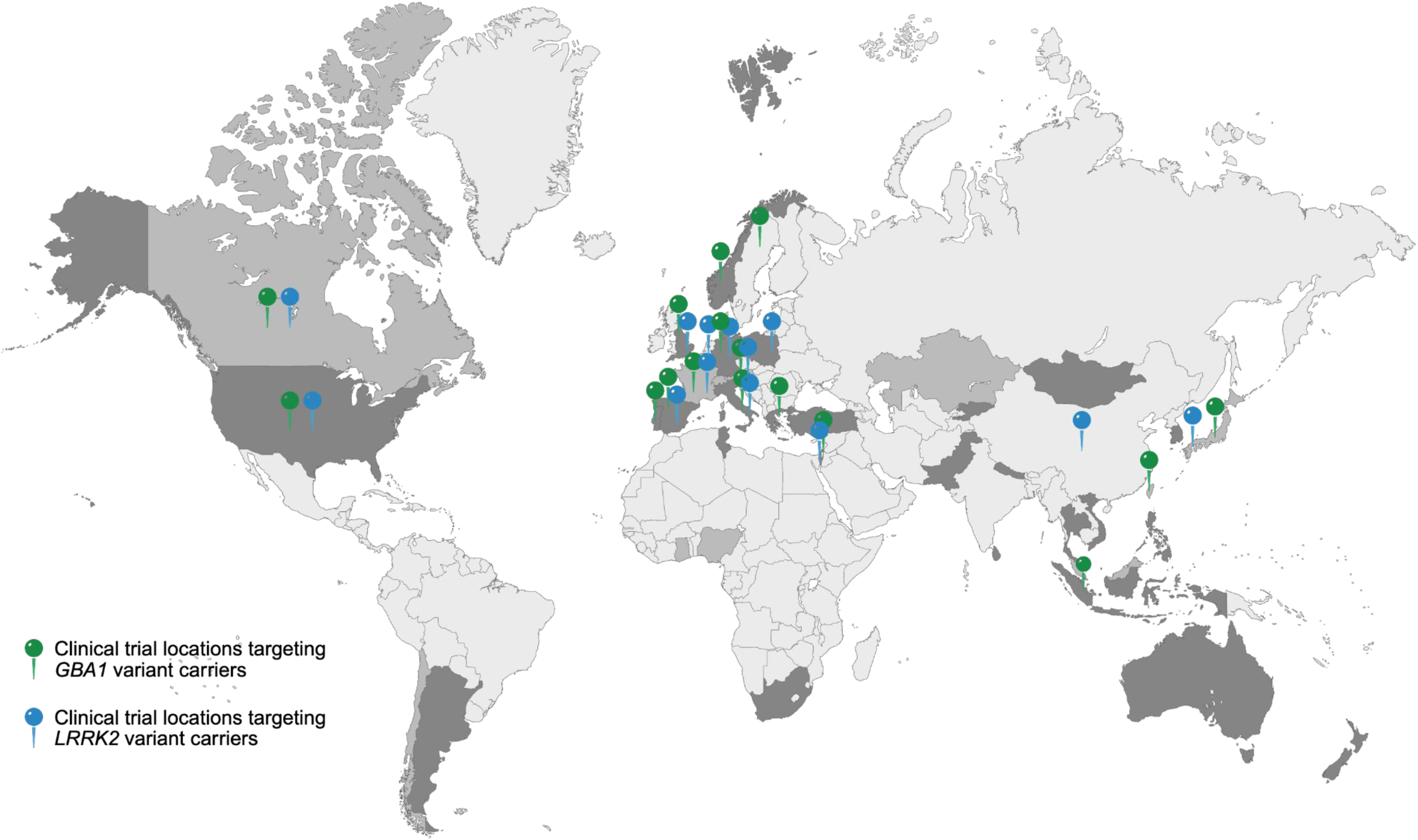
Global GP2 Precision Medicine Survey Participation, Variant Carrier Distribution, and Clinical Trial Locations. World map adapted from Lange et al., https://doi.org/10.1101/2025.07.08.25330815. Countries with centers with identified variant carriers are shown in mid-grey, and countries with centers that both participated in the survey and have identified carriers are shown in dark grey. Green and blue pins mark locations of ongoing clinical trials recruiting *GBA1* (green) and *LRRK2* (blue) variant carriers. Notably, countries are shaded once regardless of the number of participating institutions. Similarly, a single pin denotes countries with one or multiple ongoing trials.

**Figure 2.**
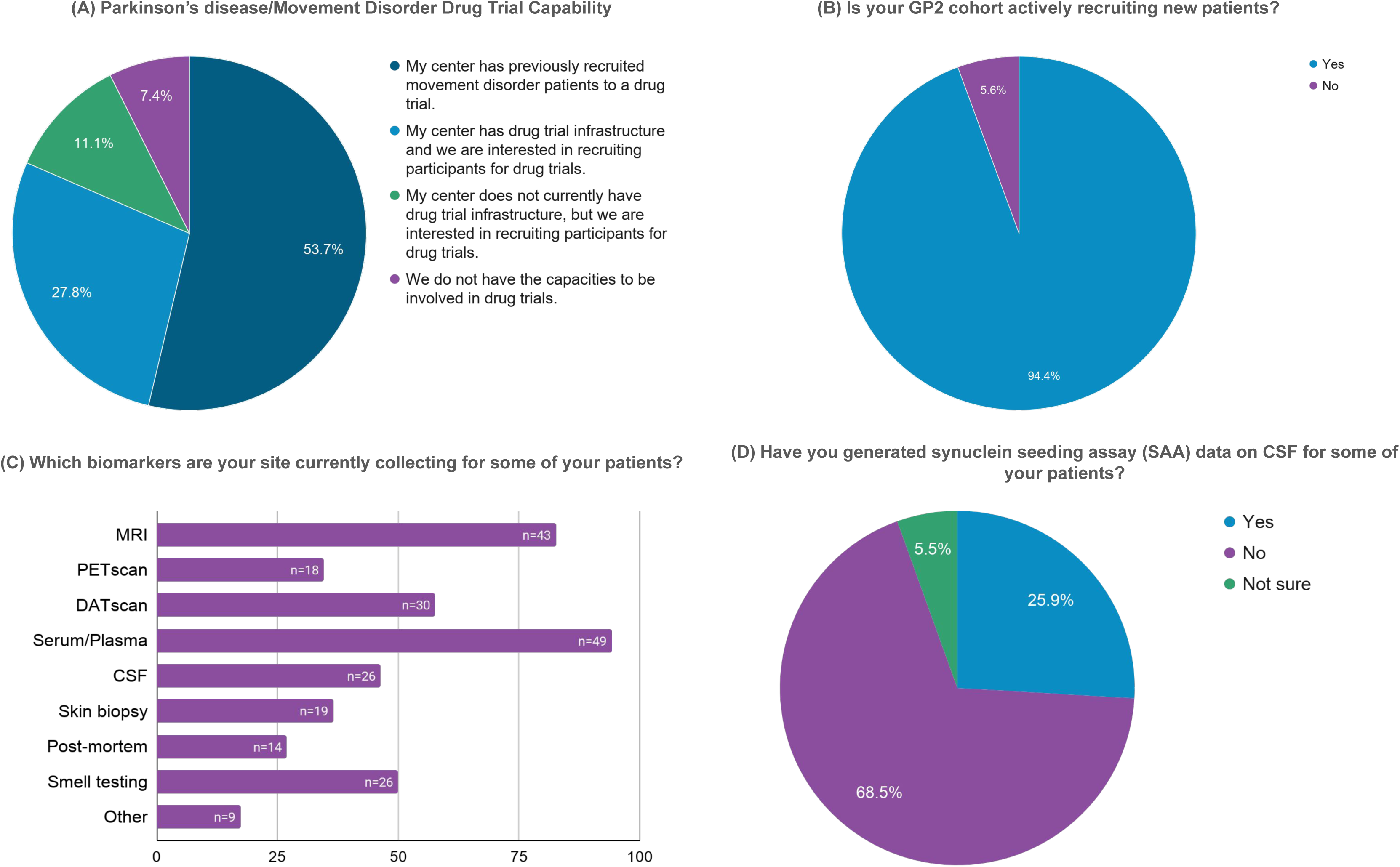
Survey responses from GP2 sites on precision medicine trial readiness, resources, and biomarker collection. Survey responses from 54 GP2 sites are summarized across pie charts (panels A, B, D) and horizontal bar charts (panel D). Panels A illustrates the sites’ capabilities to participate in PD and other movement disorder drug trials. Panel B demonstrates in GP2 sites are actively recruiting new patients. Panels C and D indicate available biomarker resources across sites. Percentages of responding centers are displayed, with absolute numbers indicated where relevant.

Out of 52 sites that currently collect different biomarkers, blood sampling for serum or plasma analyses and magnetic resonance imaging (MRI) were most frequently reported (94.2% and 82.7%, respectively). Further, DAT-SPECT imaging (57.7%), cerebrospinal fluid (CSF) collection (46.4%), or olfactory testing (50.0%) were commonly performed, while other biomarkers, including synuclein seeding assays (SAA), post-mortem brain tissue, skin biopsies, and PET imaging, were available at 25.9-36.5% of sites (**Fig. 2C and D**).

Notably, some countries in which institutions participated in the survey and had cohorts of *GBA1* and *LRRK2* variant carriers overlapped with those where ongoing GBA1- and LRRK2-targeted clinical trials are taking place, particularly across Europe (e.g., United Kingdom, Germany, Italy, Spain, Greece) and North America (e.g., the United States of America and Canada) and a few across East-Asia (e.g., Japan, Taiwan, and Indonesia). However, investigators from several regions, including South America, Africa, Asia, and Australia, reported interest and readiness despite the absence of ongoing gene-targeted trials (**Fig. 1**). Importantly, these engaged regions with limited prior trial exposure collectively account for a notable pool of genetically eligible individuals, with approximately 1,300 *GBA1* variant carriers in Africa (mostly Nigeria), ∼100 *GBA1* and ∼30 *LRRK2* variant carriers across multiple South American countries, ∼740 carriers (approximately 370 each for *GBA1* and *LRRK2* variants, respectively) across Central- and South-East-Asia (including Kazakhstan, Kyrgyzstan, Malaysia, Mongolia, Nepal, Pakistan, Turkey, and South Korea), and ∼580 *GBA1* and ∼100 *LRRK2* carriers in Australia and New Zealand.

Of note, these variant carrier counts are based on the region from which samples were submitted to GP2; however, some contributing cohorts are multi-centre, and in some instances participants or biospecimens originated from other countries (especially for larger consortia like the Parkinson’s Progression Marker Initiative [PPMI](*12*), PD GENEration(*13*), or ROPAD(*14*)). Accordingly, country-level and regional estimates should be interpreted as approximate.

## DISCUSSION

Our efforts highlight the potential value of GP2 in accelerating precision medicine initiatives for PD globally. By integrating large-scale, multi-ancestry genetic data with harmonized clinical information, GP2 provides a comprehensive, globally representative resource for identifying individuals for genotype-defined clinical trials (**Fig. 3**). Importantly, GP2 can serve as a liaison between participating clinical and research sites and pharmaceutical partners, facilitating communication and efficient recruitment of genetically characterized, potentially trial-eligible participants. The global representation is a critical first step to expanding precision medicine beyond predominantly European and North American cohorts and to ensuring equitable access to emerging gene-targeted therapies across populations.(*15, 16*)

**Figure 3.**
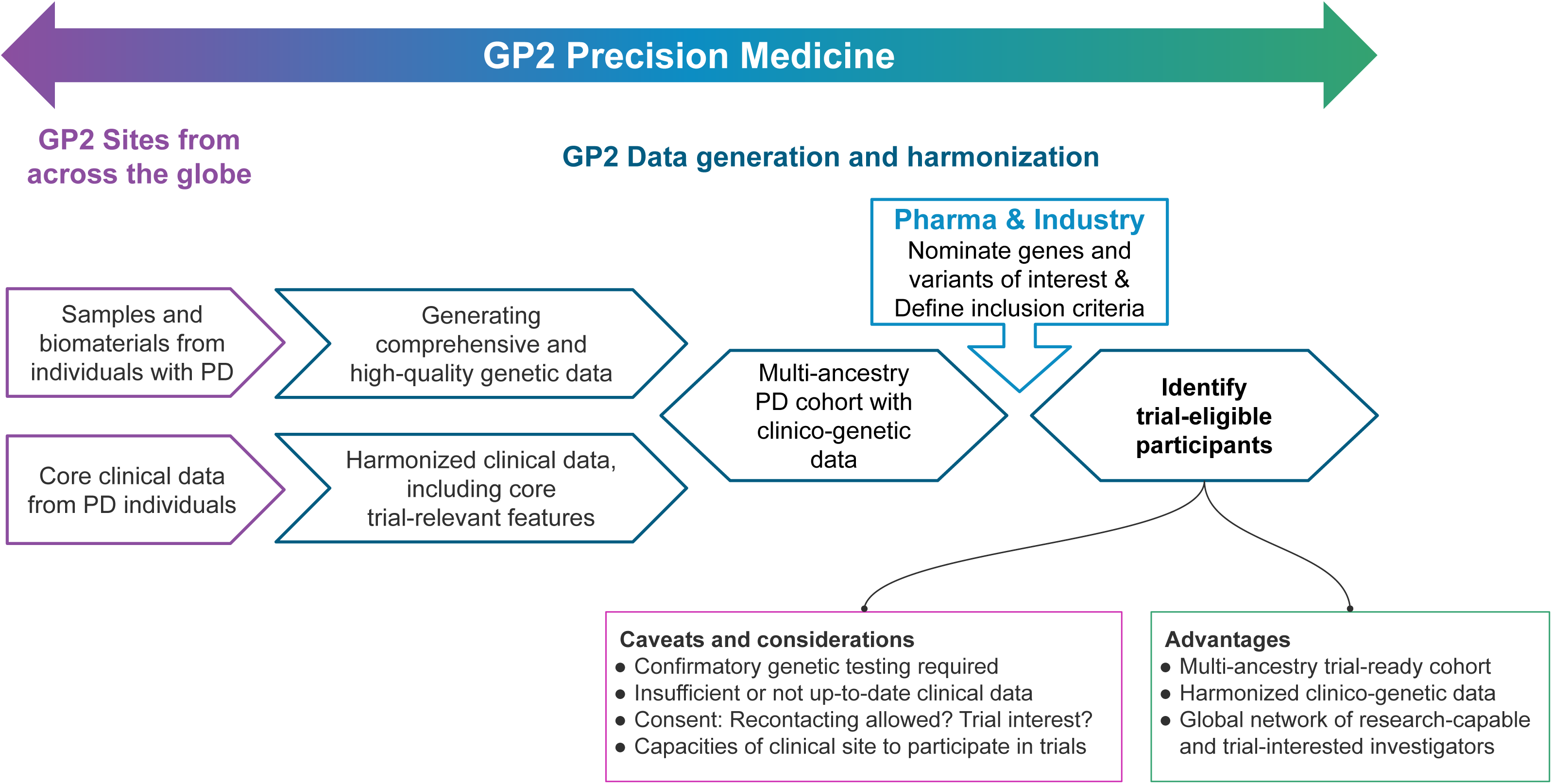
GP2 Precision Medicine Framework. GP2 sites from across the globe contribute samples and biomaterials from individuals with PD as well as corresponding clinical data on core features. By performing NBA and WGS analyses, GP2 generates comprehensive and high-quality genetic data; further, GP2 harmonizes submitted clinical data, including core trial-relevant clinical features; both together resulting in a multi-ancestry PD cohort with corresponding clinico-genetic data. Based on nominated genes or specific variants and defined inclusion criteria provided by pharma companies and industry, the GP2 PD cohort can be screened and potentially trial-eligible participants can be identified. GP2’s precision medicine group thereby functions as the liaison between PD research sites and pharmaceutical companies and industry partners.

Several ongoing clinical trials target *GBA1* and *LRRK2* variant carriers, each with distinct inclusion and exclusion criteria based on both clinical characteristics and specific genetic variants. Our exploratory assessment applied broader genetic and clinical inclusion criteria, including all carriers of confirmed pathogenic, likely pathogenic, or established risk variants, only some of which may be of interest for some trials. We focused on common clinical eligibility criteria - disease duration, disease severity, and cognitive status - while acknowledging that individual trials may include additional factors such as prior participation in interventional studies, deep brain stimulation, concomitant medications, or comorbid conditions. Furthermore, limitations in the availability or recency of clinical data, as well as varying consent structures across sites (e.g., regarding recontact or trial participation interest), may further restrict the pool of participants eligible for specific studies. Consequently, not all variant carriers identified within GP2 would meet eligibility criteria for every trial, and the number of potentially trial-eligible participants reported here likely represents an upper estimate. Nevertheless, these results highlight the practical utility of GP2 as a precision-medicine platform, as we were able to identify substantial numbers of carriers of pathogenic and high-risk *GBA1* and *LRRK2* variants across multiple ancestries and regions, providing a realistic foundation for future gene-targeted trial planning and recruitment.

Key priorities to enhance GP2’s translational impact include the systematic harmonization of clinical data and the implementation of standardized pipelines for the rapid identification and recruitment of genetically defined participants, particularly at early disease stages. Although the present analysis focused on *GBA1* and *LRRK2*, the same framework (**Fig. 3**) can be applied across all genes and variant types. Variant- and phenotype-specific inclusion criteria, defined by pharmaceutical and industry partners or emerging from research, can be systematically investigated within GP2’s integrated, multi-ancestry resource to identify individuals meeting predefined genetic and clinical criteria. Additionally, GP2 is actively collecting data on individuals with prodromal PD and family members of affected carriers, as well as healthy controls, thereby expanding the identification of non-manifesting carriers. This will accelerate recruitment into biomarker and phenoconversion studies and enable clinical trials focused on early-stage disease and prevention strategies. Notably, as a decentralized initiative, GP2 does not maintain centralized participant identifiers but instead enables controlled recontact through local study sites, thereby facilitating coordination among centers and collaborators for targeted recruitment.

Beyond providing a foundational data resource for trial recruitment, GP2 also provides a powerful platform for identifying and prioritizing new therapeutic targets. The discovery of novel risk variants, such as the African intronic *GBA1* variant rs3115534-G(*17*), the reclassification of *LRRK2* p.L1795F from a variant of uncertain significance to pathogenic(*18*), and the identification of more than 50 novel risk loci(*19*), each exemplify GP2’s ability to refine variant interpretation and expand the spectrum of actionable findings for future clinical trials. As GP2’s resources continue to grow, we anticipate that additional genes and variants of therapeutic relevance will be identified, further strengthening GP2’s role in precision medicine. Together, these efforts provide a scalable and globally representative model for linking large-scale genomic discovery with precision trial implementation in PD.

Beyond its extensive data resources, GP2 benefits from strong community engagement and international collaboration. In our survey of more than 50 participating centers worldwide, approximately three-quarters reported either prior involvement in clinical trials (∼55%) or established infrastructure and readiness to participate (∼25%), and nearly all expressed interest in partnering with pharmaceutical or industry collaborators. Moreover, nearly 90% of GP2 sites are actively recruiting participants and collecting clinical data and biomaterials. Notably, while many countries in which institutions participated in the survey also have ongoing *GBA1*- and *LRRK2*-focused clinical trials, predominantly in Europe and North America, other highly engaged regions, including South America, Africa, Asia, and Australia, currently have no such studies underway (**Fig. 1**). Together, these findings highlight the substantial capacity and commitment within the GP2 network to facilitate and accelerate precision medicine-driven clinical research, while also underscoring key geographic gaps where expanding trial availability could further enhance global equity.(*15, 16*)

Notably, as GP2 is a global research network, the participating sites included in this study represent centers with established ethical frameworks, sample collection capacity, and a minimum level of clinical and research infrastructure. As such, these findings may not fully reflect the broader landscape across all regions, particularly in underrepresented populations. In addition, while this work focused on identifying genetically defined individuals and assessing site interest and readiness, local infrastructure varies substantially, including access to specialized movement disorder care, advanced diagnostics, and biospecimen processing, which may represent a challenge for future trial development. Nevertheless, this effort provides an important first step toward establishing a global, potentially trial-eligible network and highlights key areas for future investment to support more equitable implementation of precision medicine.

Several limitations should be acknowledged. First, our analyses combined data from multiple sources (e.g., WGS, CES, and NBA), which ideally should be harmonized across individuals. This is particularly relevant given the known limitations of genotyping data for rare variant and *GBA1* analyses (e.g., certain variants are detected up by genotyping or are filtered out by the QC pipeline). Second, genetic data were generated in a research setting and will need confirmatory testing prior to trial inclusion of identified individuals. Additionally, although GP2 has significantly increased the representation of underrepresented populations in PD genetics research, the data currently collected remain disproportionately weighted toward European populations. Further, as GP2 is a continuously growing resource with ongoing clinical data collection, a considerable proportion of clinical variables essential for assessing trial eligibility remain missing for identified carriers and overall, data collection protocols differ between sites. In addition, some clinical measures used to define trial eligibility, particularly cognitive and neuropsychiatric assessments, may not be directly comparable across ancestries due to population-specific distributions, cultural factors, and differences in assessment tools. Addressing these gaps by prioritizing the systematic capture and harmonization of detailed clinical data is a major focus of ongoing GP2 efforts.

Future extensions of precision medicine may include identifying patients with specific single common variants, or polygenic risk scores interpreted in the context of ancestry-specific case-control and case-phenotype studies. It is likely that biomarker profiling will also be part of defining future trial eligibility for specific patient groups. We are well placed to contribute to these emerging efforts and initiatives.

In conclusion, leveraging its data and collaborations, GP2 directly addresses key unmet needs in PD genetics by laying the foundation for future therapeutic strategies and enabling precision medicine efforts on a global scale. Importantly, GP2’s integrated clinical and genetic framework supports the identification of trial-eligible individuals across ancestries, thereby facilitating the equitable development and recruitment of cohorts for gene carrier-targeting clinical trials.

## MATERIALS AND METHODS

### Identification of gene variant carriers and extraction of clinical data

We analyzed short-read genome (WGS) and clinical exome (CES) sequencing as well as Illumina NeuroBooster Array (NBA) (*7*) genotyping data from the latest GP2 data release (Release 11; https://doi.org/10.5281/zenodo.17753486). We included all individuals with a diagnosis of PD, including individuals with apparently idiopathic PD, individuals with an unknown but suspected genetic form of PD based on an early onset of disease (age at motor symptom onset [AAO] ≤50 years) or a positive family history of PD, and individuals with already known genetic forms of PD (genetically enriched cohorts, i.e., individuals known to carry *GBA1* or *LRRK2* pathogenic or risk variants). Deceased individuals (e.g., individuals with a provided date of death and individuals submitted from brain banks) as well as individuals with a different diagnosis (e.g., atypical or unspecified parkinsonism) were excluded. We analyzed genetic data for a total of 65,509 individuals with PD, of whom 21,748 had WGS, 14,648 had CES, and 50,073 had NBA data (for 20,684 individuals, more than one form of genetic analysis had been done).

Quality control and genetic ancestry prediction were done using GenoTools as previously described (*8*). Individuals were from 11 different genetically-determined ancestry groups: African-Admixed (AAC), African (AFR), Ashkenazi Jewish (AJ), Latinos and Indigenous People of the Americas (AMR), Complex Admixture (CAH), Central Asian (CAS), East Asian (EAS), European (EUR), Finnish (FIN), Middle Eastern (MDE), and South Asian (SAS). Notably, the CAH group comprises highly admixed individuals for whom ancestry-specific interpretation is limited. In line with the ongoing clinical trials for genetic PD, we focused our analysis on *GBA1* and *LRRK2*. Variants were included if reported as disease-causing or risk-associated in at least two of three curated databases (e.g., ClinVar, https://www.ncbi.nlm.nih.gov/clinvar/; the Human Gene Mutation Database, https://www.hgmd.cf.ac.uk; MDSGene, https://www.mdsgene.org/; **Table S1**). PLINK v.2 (*9, 10*) was used to extract variants of interest in *LRRK2* and *GBA1*, and variant annotation was done using ANNOVAR (*11*). Additionally, the Gauchian pipeline (https://github.com/Illumina/Gauchian) was used for *GBA1* variant calling from WGS data.

We reviewed eligibility criteria for PD clinical trials listed on https://clinicaltrials.gov/. Clinical variables of direct relevance to clinical trial eligibility, including disease duration, Hoehn & Yahr stage (H&Y), and Montreal Cognitive Assessment (MoCA) scores, were extracted from the most recent GP2 data release, where available. Disease duration was calculated using the age at enrollment and AAO, or age at diagnosis, if AAO was unavailable.

### Precision medicine survey of GP2 sites

We sent out a web-based survey (see Supplementary Material) to 153 GP2 sites, prioritizing those with identified variant carriers from prior exploratory analyses (excluding brain banks) as well as those with all GP2-required agreements fully executed and allowed to share data and samples with GP2. The survey was designed to evaluate their interest and capability, including available local institutional resources, to participate in precision medicine efforts and potential recruitment of individuals for clinical trials. We further gathered information on biomaterial collection at each institution.

## Acknowledgments

This project was supported by the Global Parkinson’s Genetics Program (GP2; https://gp2.org). GP2 is funded by the Aligning Science Across Parkinson’s (ASAP) initiative and implemented by The Michael J. Fox Foundation for Parkinson’s Research (MJFF). For a complete list of GP2 members see doi.org/10.5281/zenodo.7904831.

## Data and materials availability

Data used in the preparation of this article were obtained from the Global Parkinson’s Genetics Program (GP2; https://gp2.org). Specifically, we used Tier 2 data from GP2 release 11 (https://doi.org/10.5281/zenodo.17753486). Tier 1 data can be accessed by completing a form on the Accelerating Medicines Partnership in Parkinson’s Disease (AMP-PD) website (https://amp-pd.org/register-for-amp-pd). Tier 2 data access requires approval and a Data Use Agreement signed by your institution. AMP-PD data can be accessed through the AMP-PD website (https://amp-pd.org). Qualified researchers are encouraged to apply for direct access to the data through AMP-PD.

The survey data reported in this manuscript as well as all code generated for this article, and the identifiers for all software programs and packages used, are available on GitHub (https://github.com/GP2code/GP2-precision-medicine/) and were given a persistent identifier via Zenodo (https://doi.org/10.5281/zenodo.19228213).

## Funding

This research was supported in part by the Intramural Research Program of the National Institutes of Health (NIH). The contributions of the NIH author(s) are considered Works of the United States Government. The findings and conclusions presented in this paper are those of the author(s) and do not necessarily reflect the views of the NIH or the U.S. Department of Health and Human Services.

## Competing interests

KAB has received honoraria for her participation in the GBA1 Canada (G-Can) scientific advisory board. LML reports honoraria from the International Parkinson and Movement Disorder Society. ZHF, HI, LJ, HLL, DV, and MAN’s participation in this project was part of a competitive contract awarded to DataTecnica LLC by the National Institutes of Health to support open science research. IFM receives honoraria for his role in PD GENEration steering committee. CK is a medical advisor to Centogene and Biogen and has received speakers’ honoraria from Bial and royalties from Oxford University Press and Springer Nature. AJN reports grants from Parkinson’s UK, Barts Charity, Cure Parkinson’s, National Institute for Health and Care Research, UKRI, the Medical College of Saint Bartholomew’s Hospital Trust, Alchemab, and the Michael J Fox Foundation. AJN reports consultancy and personal fees from AbbVie, Bial, and Roche. AJN reports honoraria from the International Parkinson and Movement Disorders Society. MAN owns stock in Character Bio Inc and Neuron23 Inc. AS is the lead investigator for a grant from the Michael J Fox Foundation for Parkinson’s Research and has a contract for work on the Global Parkinson’s Genetics Program (GP2). He is named as an inventor on patents for a diagnostic for stroke and for molecular testing for C9orf72 repeats; he is on the scientific advisory board of the Lewy Body Disease Association (unpaid position) and for Cajal Neuroscience (paid position); and he received an honorarium for speaking at the World Laureates Association. CB is an employee of the Coalition for Aligning Science (CAS). HRM is employed by UCL. He reports paid consultancy from Arvinas, Aprinoia, Skyhawk, AI Therapeutics, Neuron23; lecture fees/honoraria - Movement Disorders Society, Bial, Calico. Research Grants from Parkinson’s UK, Cure Parkinson’s Trust, PSP Association, Medical Research Council, Michael J Fox Foundation, NIHR. HRM is a co-applicant on a patent application related to C9orf72 - Method for diagnosing a neurodegenerative disease (PCT/GB2012/052140). All other authors declare no relevant competing interests.

## Notes

### Author Declarations

This study uses data from the Global Parkinson's Genetics Program (GP2). All sites participating in GP2 and contributing samples and data to GP2 have ethical approval from their local institutions. Consent forms and ethical approvals for each participating site have been reviewed by GP2's Compliance group.

## References

1. Z. Hyderi, S. Farhana M, T. P. Singh, A. V. Ravi, Therapeutic targeting of autosomal Parkinson’s disease by modulation of leucine-Rich Repeat Kinase 2 (LRRK2) protein. Brain Res. 1860, 149674 (2025).

2. M. Karami, P. M. Sanaye, A. Ghorbani, R. Amirian, P. Goleij, M. Babamohamadi, Z. Izadi, Recent advances in targeting LRRK2 for Parkinson’s disease treatment. J. Transl. Med. 23, 754 (2025).

3. E. Menozzi, M. Toffoli, A. H. V. Schapira, Targeting the GBA1 pathway to slow Parkinson disease: Insights into clinical aspects, pathogenic mechanisms and new therapeutic avenues. Pharmacol. Ther. 246, 108419 (2023).

4. Z. Gan-Or, Lessons and future directions for GBA1-targeting therapies. Lancet Neurol. 22, 644–645 (2023).

5. E. V. Minikel, J. L. Painter, C. C. Dong, M. R. Nelson, Refining the impact of genetic evidence on clinical success. Nature 629, 624–629 (2024).

6. Global Parkinson’s Genetics Program, GP2: The Global Parkinson’s Genetics Program. Mov Disord 36, 842–851 (2021).

7. S. Bandres-Ciga, F. Faghri, E. Majounie, M. J. Koretsky, J. Kim, K. S. Levine, H. Leonard, M. B. Makarious, H. Iwaki, P. W. Crea, D. G. Hernandez, S. Arepalli, K. Billingsley, K. Lohmann, C. Klein, S. J. Lubbe, E. Jabbari, P. Saffie-Awad, D. Narendra, A. Reyes-Palomares, J. P. Quinn, C. Schulte, H. R. Morris, B. J. Traynor, S. W. Scholz, H. Houlden, J. Hardy, S. Dumanis, E. Riley, C. Blauwendraat, A. Singleton, M. Nalls, J. Jeff, D. Vitale, NeuroBooster Array: A Genome-Wide Genotyping Platform to Study Neurological Disorders Across Diverse Populations medRxiv (2023), doi:10.1101/2023.11.06.23298176.

8. D. Vitale, M. Koretsky, N. Kuznetsov, S. Hong, J. Martin, M. James, M. B. Makarious, H. Leonard, H. Iwaki, F. Faghri, C. Blauwendraat, A. B. Singleton, Y. Song, K. Levine, A. A. K. Sreelatha, Z.-H. Fang, M. Nalls, GenoTools: An Open-Source Python Package for Efficient Genotype Data Quality Control and Analysis bioRxiv (2024), doi:10.1101/2024.03.26.586362.

9. C. C. Chang, C. C. Chow, L. C. Tellier, S. Vattikuti, S. M. Purcell, J. J. Lee, Second-generation PLINK: rising to the challenge of larger and richer datasets. Gigascience 4, 7 (2015).

10. S. Purcell, B. Neale, K. Todd-Brown, L. Thomas, M. A. R. Ferreira, D. Bender, J. Maller, P. Sklar, P. I. W. de Bakker, M. J. Daly, P. C. Sham, PLINK: a tool set for whole-genome association and population-based linkage analyses. Am. J. Hum. Genet. 81, 559–575 (2007).

11. K. Wang, M. Li, H. Hakonarson, ANNOVAR: functional annotation of genetic variants from high-throughput sequencing data. Nucleic Acids Res. 38, e164 (2010).

12. Parkinson Progression Marker Initiative, The Parkinson Progression Marker Initiative (PPMI). Prog. Neurobiol. 95, 629–635 (2011).

13. L. Cook, J. Verbrugge, T.-H. Schwantes-An, J. Schulze, T. Foroud, A. Hall, K. S. Marder, I. F. Mata, N. E. Mencacci, M. A. Nance, M. A. Schwarzschild, T. Simuni, S. Bressman, A.-M. Wills, H. H. Fernandez, I. Litvan, K. E. Lyons, H. A. Shill, C. Singer, T. F. Tropea, N. Vanegas Arroyave, J. Carbonell, R. Cruz Vicioso, L. Katus, J. F. Quinn, P. D. Hodges, Y. Meng, S. P. Strom, C. Blauwendraat, K. Lohmann, C. Casaceli, S. C. Rao, K. Ghosh Galvelis, A. Naito, J. C. Beck, R. N. Alcalay, Parkinson’s disease variant detection and disclosure: PD GENEration, a North American study. Brain 147, 2668–2679 (2024).

14. A. Westenberger, V. Skrahina, T. Usnich, C. Beetz, E.-J. Vollstedt, B.-H. Laabs, J. J. Paul, F. Curado, S. Skobalj, H. Gaber, M. Olmedillas, X. Bogdanovic, N. Ameziane, N. Schell, J. O. Aasly, M. Afshari, P. Agarwal, J. Aldred, F. Alonso-Frech, R. Anderson, R. Araújo, D. Arkadir, M. Avenali, M. Balal, S. Benizri, S. Bette, P. Bhatia, M. Bonello, P. Braga-Neto, S. Brauneis, F. E. C. Cardoso, F. Cavallieri, J. Classen, L. Cohen, D. Coletta, D. Crosiers, P. Cullufi, K. Dashtipour, M. Demirkiran, P. de Carvalho Aguiar, A. De Rosa, R. Djaldetti, O. Dogu, M. G. Dos Santos Ghilardi, C. Eggers, B. Elibol, A. Ellenbogen, S. Ertan, G. Fabiani, B. H. Falkenburger, S. Farrow, T. Fay-Karmon, G. J. Ferencz, E. T. Fonoff, Y. D. Fragoso, G. Genç, A. Gorospe, F. Grandas, D. Gruber, M. Gudesblatt, T. Gurevich, J. Hagenah, H. A. Hanagasi, S. Hassin-Baer, R. A. Hauser, J. Hernández-Vara, B. Herting, V. K. Hinson, E. Hogg, M. T. Hu, E. Hummelgen, K. Hussey, J. Infante, S. H. Isaacson, S. Jauma, N. Koleva-Alazeh, G. Kuhlenbäumer, A. Kühn, I. Litvan, L. López-Manzanares, M. Luxmore, S. Manandhar, V. Marcaud, K. Markopoulou, C. Marras, M. McKenzie, M. Matarazzo, M. Merello, B. Mollenhauer, J. C. Morgan, S. Mullin, T. Musacchio, B. Myers, A. Negrotti, A. Nieves, Z. Nitsan, N. Oskooilar, Ö. Öztop-Çakmak, G. Pal, N. Pavese, A. Percesepe, T. Piccoli, C. Pinto de Souza, T. Prell, M. Pulera, J. Raw, K. Reetz, J. Reiner, D. Rosenberg, M. Ruiz-Lopez, J. Ruiz Martinez, E. Sammler, B. L. Santos-Lobato, R. Saunders-Pullman, I. Schlesinger, C. M. Schofield, A. F. Schumacher-Schuh, B. Scott, Á. Sesar, S. J. Shafer, R. Sheridan, M. Silverdale, R. Sophia, M. Spitz, P. Stathis, F. Stocchi, M. Tagliati, Y. F. Tai, A. Terwecoren, S. Thonke, L. Tönges, G. Toschi, V. Tumas, P. P. Urban, L. Vacca, W. Vandenberghe, E. M. Valente, F. Valzania, L. Vela-Desojo, C. Weill, D. Weise, J. Wojcieszek, M. Wolz, G. Yahalom, G. Yalcin-Cakmakli, S. Zittel, Y. Zlotnik, K. K. Kandaswamy, A. Balck, H. Hanssen, M. Borsche, L. M. Lange, I. Csoti, K. Lohmann, M. Kasten, N. Brüggemann, A. Rolfs, C. Klein, P. Bauer, Relevance of genetic testing in the gene-targeted trial era: the Rostock Parkinson’s disease study. Brain 147, 2652–2667 (2024).

15. S.-Y. Lim, A. H. Tan, A. Ahmad-Annuar, N. U. Okubadejo, K. Lohmann, H. R. Morris, T. S. Toh, Y. W. Tay, L. M. Lange, S. Bandres-Ciga, I. Mata, J. N. Foo, E. Sammler, J. C. E. Ooi, A. J. Noyce, N. Bahr, W. Luo, R. Ojha, A. B. Singleton, C. Blauwendraat, C. Klein, Uncovering the genetic basis of Parkinson’s disease globally: from discoveries to the clinic. Lancet Neurol. 23, 1267–1280 (2024).

16. L. M. Lange, Z.-H. Fang, M. B. Makarious, N. Kuznetsov, K. A. Brolin, S. Ballard, S. Bardien, M. L. Doquenia, P. Heutink, H. Houlden, H. Iwaki, S. Jasaityte, L. Jones, J. Junker, R. Kaiyrzhanov, M. J. Koretsky, K. R. Kumar, Latin American Research Consortium on the Genetics of Parkinson’s Disease (LARGE-PD), H. L. Leonard, K. S. Levine, S.-Y. Lim, N. E. Mencacci, W. M. Y. Mohamed, M. A. Nalls, A. J. Noyce, R. Ojha, N. U. Okubadejo, S. U. Rehman, L. Screven, C. Shashkin, S. Sopromadze, E. J. Stafford, A. H. Tan, M. Tan, Z. Tavadyan, J. Trinh, B. Tserensodnom, E. M. Valente, D. Vitale, N. Zharkinbekova, K. Lohmann, S. Bandres-Ciga, C. Blauwendraat, A. Singleton, H. R. Morris, C. Klein, Global Parkinson’s Genetics Program (GP2), The global landscape of genetic variation in Parkinson’s disease: Multi-ancestry insights into established disease genes and their translational relevance medRxiv (2025), doi:10.1101/2025.07.08.25330815.

17. M. Rizig, S. Bandres-Ciga, M. B. Makarious, O. O. Ojo, P. W. Crea, O. V. Abiodun, K. S. Levine, S. A. Abubakar, C. O. Achoru, D. Vitale, O. A. Adeniji, O. P. Agabi, M. J. Koretsky, U. Agulanna, D. A. Hall, R. O. Akinyemi, T. Xie, M. W. Ali, E. A. Shamim, I. Ani-Osheku, M. Padmanaban, O. M. Arigbodi, D. G. Standaert, A. H. Bello, M. N. Dean, C. O. Erameh, I. Elsayed, T. H. Farombi, O. Okunoye, M. B. Fawale, K. J. Billingsley, F. A. Imarhiagbe, P. A. Jerez, E. U. Iwuozo, B. Baker, M. A. Komolafe, L. Malik, P. O. Nwani, K. Daida, E. O. Nwazor, A. Miano-Burkhardt, Y. W. Nyandaiti, Z.-H. Fang, Y. O. Obiabo, J. H. Kluss, O. A. Odeniyi, D. G. Hernandez, F. E. Odiase, N. Tayebi, F. I. Ojini, E. Sidranksy, G. A. Onwuegbuzie, A. M. D’Souza, G. O. Osaigbovo, B. Berhe, N. Osemwegie, X. Reed, O. O. Oshinaike, H. L. Leonard, F. M. Otubogun, C. X. Alvarado, S. I. Oyakhire, S. I. Ozomma, S. C. Samuel, F. T. Taiwo, K. W. Wahab, Y. A. Zubair, H. Iwaki, J. J. Kim, H. R. Morris, J. Hardy, M. A. Nalls, K. Heilbron, L. Norcliffe-Kaufmann, Nigeria Parkinson Disease Research Network, International Parkinson’s Disease Genomics Consortium Africa, Black and African American Connections to Parkinson’s Disease Study Group, 23andMe Research Team, C. Blauwendraat, H. Houlden, A. Singleton, N. U. Okubadejo, Global Parkinson’s Genetics Program, Identification of genetic risk loci and causal insights associated with Parkinson’s disease in African and African admixed populations: a genome-wide association study. Lancet Neurol. 22, 1015–1025 (2023).

18. L. M. Lange, K. Levine, S. H. Fox, C. Marras, N. Ahmed, N. Kuznetsov, D. Vitale, H. Iwaki, K. Lohmann, L. Marsili, A. J. Espay, P. Bauer, C. Beetz, J. Martin, S. A. Factor, L. A. Higginbotham, H. Chen, H. Leonard, M. A. Nalls, N. E. Mencacci, H. R. Morris, A. B. Singleton, C. Klein, C. Blauwendraat, Z.-H. Fang, Global Parkinson’s Genetics Program (GP2), The LRRK2 p.L1795F variant causes Parkinson’s disease in the European population. NPJ Parkinsons Dis. 11, 58 (2025).

19. H. L. Leonard, Global Parkinson’s Genetics Program (GP2), Novel Parkinson’s disease genetic risk factors within and across European populations. medRxiv (2025), doi:10.1101/2025.03.14.24319455.

