## Supplementary Material for "Bridging Genetics and Precision Medicine in Parkinson’s Disease through GP2"

^†^ Shared first authors.

^‡^ Joint last authors.

### **Survey questionnaire**

1. Parkinson’s disease or other movement disorder drug trials capabilities
   a. My center has previously recruited movement disorder patients to a drug trial.
   b. My center has drug trial infrastructure and we are interested in recruiting participants for drug trials.
   c. My center does not currently have drug trial infrastructure, but we are interested in recruiting participants for drug trials.
   d. We do not have the capacity to be involved in drug trials.
2. Has your site been or do you plan to be involved in an interventional clinical trial for any Parkinsonisn condition between 01/01/2022 and 01/01/2026?
   a. Yes
   b. No
   c. Not sure
3. Please indicate your study code or the name of your GP2 site.
   a. Free text
4. Is your GP2 cohort a single center or a multi-center cohort?
   a. Single center
   b. Multi-center
5. Is your cohort actively recruiting new patients?
   a. Yes
   b. No
6. Which clinical trials for PD or other movement disorders is your site currently participating in?
   a. Free text
7. Would you be happy for GP2 to connect you with pharmaceutical companies regarding potentially trial-eligible participants from your study?
   a. Yes
   b. No
8. Which biomarkers are your site currently collecting for some of your patients?
   a. MRI
   b. PETscan
   c. DATscan
   d. Blood for plasma/serum
   e. CSF
   f. Skin biopsy
   g. Post-mortem brain donation
   h. Smell testing
   i. Other (free text)
9. Which biomarkers would you possibly be able to collect for some of your patients?
   a. MRI
   b. PETscan
   c. DATscan
   d. Blood for plasma/serum
   e. CSF
   f. Skin biopsy
   g. Post-mortem brain donation
   h. Smell testing
   i. Other (free text)
10. Have you generated synuclein seeding assay (SAA) data on CSF for some of your patients?
    a. Yes
    b. No
    c. Unsure
